# Parental perspectives on the grief and support needs of children and young people bereaved during the Covid-19 pandemic: Qualitative findings from a national survey

**DOI:** 10.1101/2021.12.06.21267238

**Authors:** Emily Harrop, Silvia Goss, Mirella Longo, Kathy Seddon, Anna Torrens-Burton, Eileen Sutton, Damian JJ Farnell, Alison Penny, Annmarie Nelson, Anthony Byrne, Lucy E. Selman

**Affiliations:** Cardiff University, Marie Curie Research Centre, Division of Population Medicine, Cardiff, UK; Cardiff University, PRIME Centre, Division of Population Medicine, Cardiff, UK; University of Bristol, Palliative and End of Life Care Research Group, Population Health Sciences, Bristol Medical School, Bristol, UK; Cardiff University, School of Dentistry, Cardiff, UK; National Bereavement Alliance/Childhood Bereavement Network, London, UK

**Keywords:** Bereavement, Grief, Pandemics, Coronavirus Infections, Bereavement Services, Children, Young People

## Abstract

**Background:** During the Covid-19 pandemic, many children and young people have experienced the death of close family members, whilst also facing unprecedented disruption to their lives. This study aimed to investigate the experiences and support needs of bereaved children and young people from the perspective of their parents and guardians.

**Methods:** We analysed cross-sectional qualitative free-text data from a survey of adults bereaved in the UK during the pandemic. Participants were recruited via media, social media, national associations and community/charitable organisations. Thematic analysis was conducted on free text data collected from parent/guardian participants in response to a survey question on the bereavement experiences and support needs of their children.

**Results:** Free-text data from 104 parent and grandparent participants was included. Three main themes were identified: the pandemic-related challenges and struggles experienced by children and young people; family support and coping; and support from schools and services. Pandemic-related challenges include the impacts of being separated from the relative prior to their death, isolation from peers and other family members, and disruption to daily routines and wider support networks. Examples were given of effective family coping and communication, but also of difficulties relating to parental grief and children’s existing mental health problems. Schools and bereavement organisations’ provision of specialist support was valued, but there was evidence of unmet need, with some participants reporting a lack of access to specialist grief or mental health support.

**Conclusion:** Children and young people have faced additional strains and challenges associated with pandemic bereavement. We recommend resources and initiatives that facilitate supportive communication within family and school settings, adequate resourcing of school and community-based specialist bereavement/mental health services, and increased information and signposting to the support that is available.

## Background

The Covid-19 pandemic has resulted in widespread global mass bereavement. Although children and young people are far less likely to suffer serious illness or death as a result of Covid-19, many have been bereaved of a close family member as a direct consequence of the virus, including 12,300 children bereaved of primary or secondary resident caregivers in the UK (e.g. parents or grandparents who live with them) (1). Children and young people bereaved due to other causes of death have also been left grappling with their loss at a time of unprecedented social disruption.

Research has shown that childhood bereavement increases the risk of adverse outcomes across the life-course (2-5). In pre-pandemic times it is estimated that 5 to 20% of bereaved children and adolescents develop psychiatric difficulties (6,7), with predictions that this may increase during the pandemic (8,9), along with psycho-social, educational and workplace problems (9). During Covid-19 it has been suggested that children may be at enhanced risk for negative mental health impacts, given their limited capacity to fully understand their surroundings and control their environments (8), and the known effects of loneliness and social isolation on depression and anxiety amongst children and adolescents (10). Various Covid-19-related risk factors for bereaved children and young people have been suggested, including bereavement after a sudden death, being unable to visit their dying family members, missing the funeral, and fear of further deaths within the family, whilst exposure to media reporting and other social abnormalities such as mask-wearing are constant reminders of their loss (8, 11, 12).

High-level disruption to daily routines, relationships and support networks, especially during periods of prolonged school closure and lockdowns, are also likely to undermine the usual coping resources and mechanisms available to children and adolescents, including the possibility of respite from their grief (8,12). Strain on family life due to work or financial pressures, combined with parental grief, may also mean that is harder for parents to provide the emotional support that young people need, whilst perceived distress or vulnerability in the parents may also stop children from talking about their feelings (8,12,13). Pre-pandemic and pandemic research has also demonstrated maladaptive tendencies amongst adults to try to protect children by not talking about the death pre- and post-bereavement (8,14-17), uncertainty about how best to prepare children for the death (15,18), and difficulties maintaining parenting roles in the midst of their own grief and disruption (4). The difficulties of bereaved children and young people in managing and expressing their feelings has also been documented (4). Clear and honest communication with children about the death has been demonstrated to improve psychosocial, mental and physical health outcomes (19,20).

Having the right support in place for bereaved children and young people is essential for promoting resilience and mitigating the risks of future mental health and other problems (21). As in public health models for adult bereavement care (22), a tiered approach has been conceptualised for children and young people. According to this model, all children, young people and their parents or carers should receive information on how children grieve, what can help, and when and where to seek help, combined with a supportive response from existing networks. Some will also need support from services specialising in child bereavement, whilst the small proportion of those deemed vulnerable or traumatised will require individual or family intervention from specialist mental health services (23). Schools and other community services have a role to play in providing information on grief symptoms and informally supporting children and families known to be bereaved, as well as assessing and addressing student needs for more formal emotional support (8,24,25,26).

Evidence on adult bereavement during the pandemic is growing (27-30), however there is a lack of research relating to bereaved children and young people, specifically in relation to their grief experiences, support networks and access to specialist support services. Based on free-text data from a national survey of adults bereaved in the UK during COVID-19, this paper reports parent/guardian perspectives on their children’s bereavement support needs and experiences. Policy and practice implications are identified for improving the support for children, young people and their families during and beyond the Covid-19 crisis.

## Methods

### Study design and aim

Free-text results are reported from the second round of a longitudinal survey which aims to investigate the grief experiences, support needs and use of bereavement support by people bereaved during the pandemic. The Checklist for Reporting Results of Internet E-Surveys (31) was followed.

### Survey development

An open web survey was designed by the research team, which includes a public representative (KS), with input from the study advisory group. It was piloted, refined with public representatives with experience of bereavement and tested by the study advisory group and colleagues. Open and closed questions covered grief experiences, and perceived needs for, access to and experiences of formal and informal bereavement support (27). In response to stakeholder requests, the second round survey included a question which asked about children or young people living with the respondent, including their ages, and the free text question ’Please tell us about any support that you feel they need and/or any support they have been receiving’ (supplementary file one). These data have been analysed for this paper.

### Study procedure

The baseline survey was administered via JISC (https://www.onlinesurveys.ac.uk/) and was open from 28th August 2020 to 5th January 2021 (27,28). It was disseminated to a convenience sample from social and mainstream media and via voluntary sector associations and bereavement support organisations, including those working with ethnic minority communities. Organisations helped disseminate the voluntary (non-incentivised) survey by sharing on social media, web-pages, newsletters, on-line forums and via direct invitations to potential participants. For ease of access, the survey was posted onto a bespoke study-specific website with a memorable URL (www.covidbereavement.com) (27,28). The second follow up survey reported here was sent to baseline participants who consented to receive follow up surveys around seven months post date of death. These were personalised for each participant using individual survey links, labelled with their participant study IDs. Where baseline surveys were completed at least five months post-death (or the date of death was not given), the second survey was sent out two months after the first survey was received. All second round surveys were completed between 20/11/20 and 24/08/2021 and on average 242 days (median = 234 days or 8 months) after the date of death (range 145 to 345 days).

Inclusion criteria for study enrolment: aged 18+; family member or close friend bereaved since social-distancing requirements were introduced in the UK (16/03/2020); death occurred in the UK; ability to consent. The initial section of the survey requested informed consent and details data protection.

An additional inclusion criterion for this analysis was that participants reported on the experiences and/or needs of at least one child or young person aged 25 or under who lived with them.

### Data analysis

Free-text survey responses were analysed using inductive thematic analysis, involving line-by-line coding in Excel and identification of descriptive and analytical themes (32). The framework was revised and applied in an iterative process moving between the data and the analytical concepts to develop codes and themes grounded in the data. This involved double coding of the data set (EH, SG) and discussion and review of final themes by the research team. 27% (n=104) of second round participants (n=384) provided comments that related to their child/children’s bereavement experiences and support needs.

## Results

### Participants

Responses were included from 104 participants. Two participants were grandparents and the remainder were parents (n=102). Most deaths were caused by Covid-19 (n=55), followed by cancer (n=21). The total number of children or young people reported was 176, with a median age of 13 (range 2-25 years). The children and young people in these families were most commonly bereaved of grandparents (n=81), followed by parents (n=14), siblings (n=2) and aunts/uncles (n=2) (Table 1).

**Table 1:** participant characteristics (n=104)

|  | *N* (%) |
| --- | --- |
| **Gender of adult participant** |  |
| Male | 7 (6.7) |
| Female | 97 (93.3) |
| **Ethnicity of adult participant** |  |
| White English | 15 (14.4) |
| White British | 69 (66.3) |
| White Northern Irish | 1 (1.0) |
| White Scottish | 8 (7.7) |
| White Welsh | 7 (6.7) |
| White Australian | 1 (1.0) |
| White and Asian | 3 (2.9) |
| **Relationship of the person who died to the adult participant** |  |
| Their child | 3 (2.9) |
| Their husband/male partner | 12 (11.5) |
| Their wife/female partner | 2 (1.9) |
| Their parent(s) | 80 (76.9) |
| Their siblings | 2 (1.9) |
| Their father-in-law/step-father | 4 (3.8) |
| Their grandparent | 1 (1.0) |
| **Relationship of the person who died to the child/young person** |  |
| Their parents | 14 (13.5) |
| Their grandparents | 81 (77.9) |
| Their siblings | 2 (1.9) |
| Their Aunts/Uncles | 2 (1.9) |
| Their Great-grandparents | 1 (1.0) |
| Other family members | 4 (3.8) |
| **Cause of death** |  |
| Confirmed/suspected Covid-19 | 55 (52.9) |
| Cancer | 21 (20.2) |
| Other | 27 (25.9) |
| Don't know | 1 (1.0) |
|  | Age in years |
| **Age of children (n=176)** |  |
| Mean [Median] | 13.1 [13] |
| Range | 2-25 |

### Themes

Three main themes and eleven sub-themes (Table 2) were identified: Challenges and struggles; family support and coping; and support from schools and services. Where observed, group differences such as those relating to the age of the child, or their relationship to the person who died, are noted in the descriptive narrative.

**Table 2:**
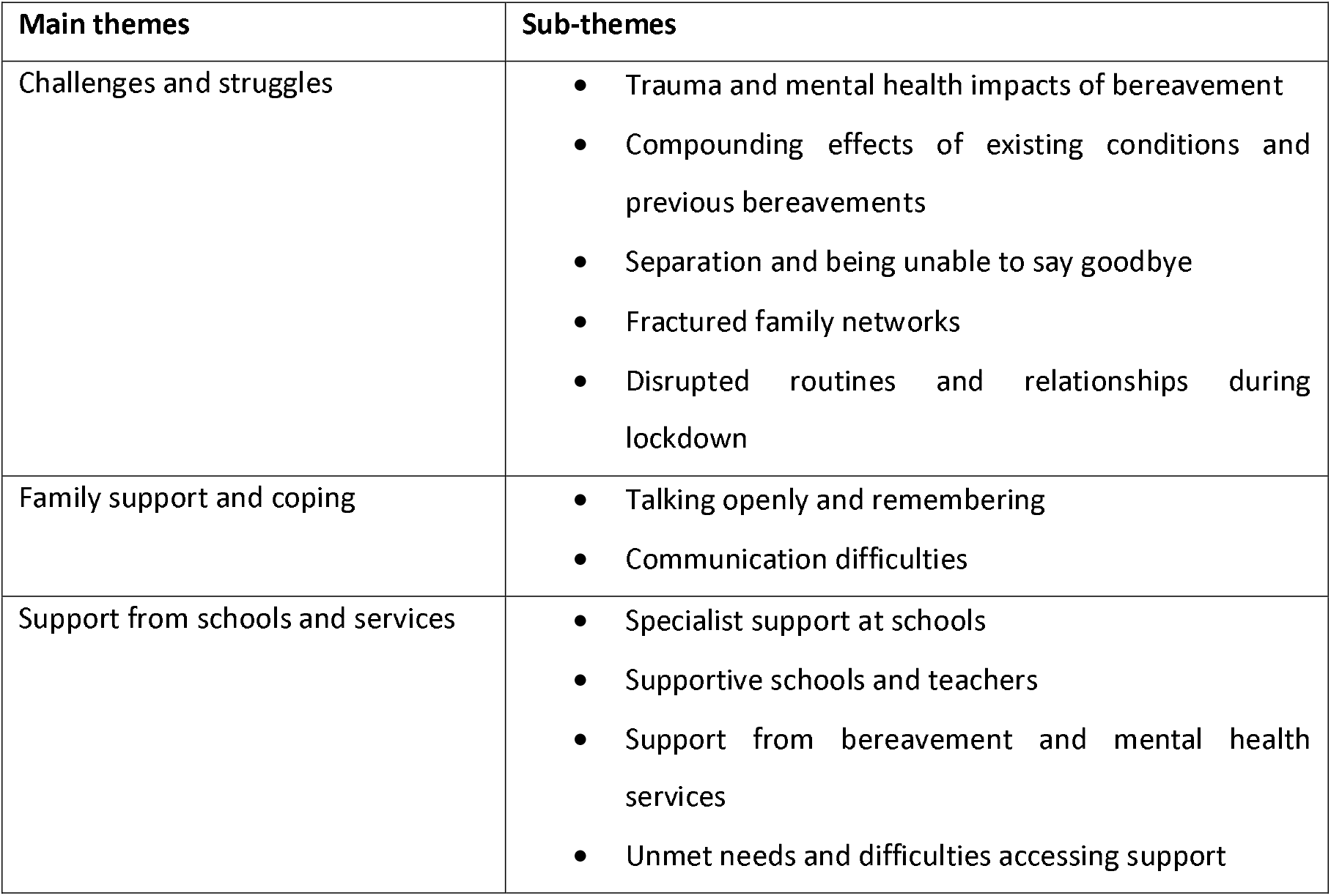
Themes.

| Main themes | Sub-themes |
| --- | --- |
| Challenges and struggles | • Trauma and mental health impacts of bereavement<br>• Compounding effects of existing conditions and previous bereavements<br>• Separation and being unable to say goodbye<br>• Fractured family networks<br>• Disrupted routines and relationships during lockdown |
| Family support and coping | • Talking openly and remembering<br>• Communication difficulties |
| Support from schools and services | • Specialist support at schools<br>• Supportive schools and teachers<br>• Support from bereavement and mental health services<br>• Unmet needs and difficulties accessing support |

### Challenges and struggles

Although many respondents felt that their children were coping well, others described their children’s struggles to come to terms with the death of a parent, grandparent and other family member, with a small minority reporting signs of associated mental health problems such as night terrors, anxiety and depression. Some also described the compounding effects of the Covid bereavement experience on pre-existing mental health issues (for the child and other family members), chronic conditions and educational needs, as well as other recent close bereavements.

> ‘My [child] is struggling with her grief, which has escalated over the last month or so. My [child] also has additional support needs which makes processing the situation even more difficult for her. My [parent] was shielding for several months and then to my [child] and [other relative] … just disappeared which has been extremely difficult for everyone.’ (RID025, parent of child/children aged under 12)

As seen in the above extracts, pandemic-related issues reported to exacerbate their child’s grief included being separated from their grandparent before their death due to shielding requirements and being unable to visit them prior to their death. This separation and sense of ’disappearance’ was felt to have made it harder for them to understand and process the situation, especially when they were then also isolated from their wider family and their other grandparent due to lockdown conditions.

> ‘It would have helped them if we could have spent more time with the wider family. They miss their [grandparent] and they haven’t been able to see their [other grandparent who] is in a support bubble with my [sibling].’ (RID692, parent of child/children aged under 12 and teenage)

Respondents described the added strain caused to their children by periods of national lockdown - including school and university closures - and the resulting disruption to their child’s daily routines and relationships. Older children and young people were particularly affected by isolation from their peers, experiencing difficulties expressing their feelings and interacting with friends remotely and, in the case of one University student, having ’too much time on his hands’.

> ‘My [child] has really needed the support of […] friends which is not allowed under current Covid restrictions. [Child] has started 6th form but is unable to mingle socially so has kept [their] feelings to [themselves]. [They] had a few private therapy sessions over the summer.’ (RID030, parent of teenage child/children)

In the case of a younger child whose grandparent died of Covid-19, the parent described her child’s anxiety over further deaths and perceived responsibility of avoiding contact with the virus, manifesting in obsessive behaviours.

> ‘My [children] live at home my youngest was very close to my [parent] and was very upset when [their grandparent] died. [Child] was very nervous going back to school and then when […] was told [that they] had to isolate because they had a case in their year [the child] wouldn’t come near me and kept washing his hands in a very ’OCD” way. I asked why and [they] said ’I don’t want to give it to you […] and for you to die like [grandparent]’ but […] has been better since I do reassure [them] the best I can all the time.’ (RID595, parent of children ranging from under 12 to early 20s)

#### Family support and coping

Many participants described how they were supporting their children to manage and cope with their bereavement. They described how they tried to talk openly with their children about their grief, feelings and memories of their relative, taking time to actively remember them, and in one case creating a memory garden which they could ’nourish’ together. One parent explained how they positively reflected with their children on how their grandparent escaped the further trauma of living through the pandemic. Some also reported using web-resources and books and stories relating to bereavement with their children. One grandparent noted the benefits of having a supportive network of family involved in supporting their grandchild following a parental death.

> ‘[We] have spent a lot of time with my [child] talking about my [parent] who [child] was very close to. We have looked at lots of photos […] and watched some film clips too. Our [child] has also now reached out to his other grandparent to establish a stronger relationship with [them]. [This grandparent] lives abroad but they zoom each week. I did look at the website of the charity [charity website] to give me ideas of how to support my [child].’ (RID511, parent of teenage child/children)

Several parents described how their children had chosen not to take up external support, preferring instead to talk with themselves and other friends and family. This was felt to be adequate in most cases, but some described their children struggling to open up to them, as well as their own challenges providing this support when coping with their own grief, perceiving a need for help to enable them to better support their child. Several parents described how their children worried when they (the parents) became upset, describing how they tried to stay strong to help protect their children.

> ‘I think my [child] need support but he doesn’t want to speak to a counsellor. [They] sometimes speaks to me and gets anxious and stressed but I don’t always feel I have the emotional resilience to cope and this makes me feel terrible.’ (RID518, Parent of teenage child/children)

#### Support from schools and services

Just under a quarter (n=19) of respondents who had school-aged children described support received through schools following their bereavements. In roughly half of these cases additional, specialist emotional support had been received, including counselling, drama therapy and support from school-based pastoral teams and emotional literacy support assistants (ELSA). These interventions were valued not just for the help already given to their children, but also knowing that it was there if needed again in the future. However, it was noted how some of these services stopped during periods of school closure, with other programmes not available at all during the pandemic.

> ‘My [child] received some counselling at school. [Their] teachers have also been very supportive. […] only had a few counselling sessions as [they] felt that [they] didn’t need or want more. [They] know that the service is there though’ (RID394, parent of teenage child/children)

Others described more general, but highly valued support provided by schools and teachers; only one parent described a lack of support and understanding. Features of valued support included checking in on students during closure periods, being aware of the student’s bereavement circumstances and potential problems, and proactively offering or placing students on the ’radar’ for specialist emotional support if needed. Some also described the skills of teachers, trained in bereavement support, who discussed the bereavement with their children and themselves, supporting not only the children in their grief, but also the parent in tending to their child’s needs.

> ‘Both my [children] have been very well supported by their school. Their teachers have not shied away from having difficult conversations with them, and have supported them in their grief.’ (RID574, parent of child/children aged under 12)

Just under a quarter of respondents (n=24) described needing or receiving additional support from bereavement or mental health services, for at least one of their children. Nine reported needing (but not receiving) additional support from bereavement or mental health services, almost all relating to loss of grandparents to Covid-19. Reasons for needing and receiving specialist support included existing mental health conditions exacerbated by bereavement (amongst older children), as well as bereavement-associated trauma and distress relating to sudden, mostly Covid-19 deaths and a lack of understanding about what happened. Parents also described their children’s needs to talk to someone outside of the family about their feelings, their struggles coming to terms with the loss of a particularly close grandparent, other recent bereavements and first experiences of death, and the added complexities caused by not having been able to say goodbye and the effects of social isolation.

> ‘Need for support to talk to someone other than family about how they are feeling. Was tough as at the time of the bereavement as they had no face-to-face contact with friends and less able to talk about their feelings.’ (RID040, parent of teenage child/children)

The counselling and psychological support received was mostly found to be helpful, with the positive relationships and skills of therapists/counsellors noted. However, a couple of parents felt that their children needed more sessions than the allocated amount, noting how they could not afford to pay for private sessions, as well as their child’s concerns about having to connect and repeat the process with a different counsellor.

> ‘They have received counselling from [children’s bereavement charity]. The counsellor is fantastic but they are only able to offer 6 - 8 weeks of 1hr sessions. This is not enough for them. My [child] is very upset that the counselling had to stop and that she will now need to go through it all again if we start a different counsellor. I can’t really afford to pay for [several] people to have counselling at once.’ (RID055 parent of teenage child/children)

One parent described difficulties with the family-based support provided, as her children were at different stages in their grief, instead perceiving a need for separate child-focused services for her youngest child.

> ‘They definitely need support as they are at very different stages. We had a family bereavement session but the younger [child] hated it [and] needs children’s services. Need to look at local groups. My older [child] was offered counselling via uni but at the time she was OK. I think now she needs to start this - I have mentioned it to [this child]. We were initially waiting to see if they could have face to face support but obviously that isn’t possible so I think online will work for now.’ (RID084 parent of teenage child/children)

Reasons why some children and young people were not getting the support needed included absence of or delayed referrals to child bereavement or mental health services caused by the pandemic, long waiting times for support, not knowing how to get support, preferences for face-to-face support, and resistance from their children to receiving external support.

> ‘[Child] has been supported by me but unable to see family as they do not live locally [and is] unable to see friends face to face - so been worse for [them]. I provide [their] bereavement support as has not wanted to use services as [child] is very sceptical that they will offer [them] anything’. (RID341 parent of teenage child/children)

## Discussion

This study describes the support needs and experiences of children and young people bereaved during the Covid-19 pandemic, as perceived by their parents or carers. Key findings include pandemic-related challenges relating to separation from the person who died prior to their death, isolation from peers and other family members and disruption to daily routines, and wider support networks during periods of school/university closure. Examples were given of effective family coping and communication, but also struggles relating to parental grief and pre-existing child mental health problems. The role of schools and bereavement charities in providing specialist support was demonstrated and valued, but there was evidence of unmet needs and lack of access to specialist grief or mental health support.

In terms of the bereavement challenges and difficulties experienced by children and young people, these findings point to the compounding effects of pre-existing mental health conditions and other recent close bereavements, as well as previously observed communication and emotional difficulties experienced by parents grappling with their own grief whilst supporting their children (8,14,15,17). They also provide empirical evidence for pandemic-specific stressors hypothesised for children and young people (8,9), and observed for bereaved adults (27-30). These include loss-related challenges such as separation from grandparents due to shielding, being unable to visit or say good-bye to them immediately prior to the death (28), and being apart from other family members in early bereavement (27,28); all factors which were felt to make the death harder to process and accept. The coping abilities of children and young people were also challenged by lack of social support caused by isolation from peers and other family members during lockdown, loss of routine, potential respite and distraction at school/ university during periods of closure, and anxiety relating to contracting and spreading the virus to other family members when schools reopened.

In terms of support, a number of family coping and communication strategies were identified, some of which are known from previous research to support healthy adaptation and grieving (8,19,20). These included open communication with children about the death and their feelings about it, actively remembering the person who died, using on-line resources, and reading books relating to grief and bereavement with their children. The important role of schools in providing informal and formal emotional support was also demonstrated, in line with previous research and established models of good practice (8,24-27). Checking in during lockdowns, being aware of the student’s bereavement circumstances, and proactively monitoring and offering students specialist emotional support if felt to be needed were valued approaches, as were the conversational skills and bereavement competencies of teachers. However, school closures during lockdowns meant that some children in need of additional supports from teachers and bereavement specialists were unable to access it when they needed it.

Although a good proportion of respondents felt that their children were coping well, findings also suggest reasonably high levels of need for specialist bereavement or mental health support amongst some of the children of our parent/caregiver sample. Around a quarter of respondents described receiving or needing additional support from either schools, bereavement charities or mental health services for at least one of their children. This need for help followed the loss of grandparents as well as parents, with unmet need most evident in relation to the death of grandparents due to Covid-19. Where children and young people were not getting the additional support that their parents perceived them to need (approximately a third of this group), this was for reasons such as pandemic-related delays to referrals, waiting lists, lack of appropriate support, concerns over the costs of obtaining private counselling support, not knowing how to get support, and resistance or disinterest from the child or young person. Recent findings from the UK that only a tenth of relatives bereaved during the pandemic were asked about their deceased relative’s relationships with children (15), and only a third were provided with information about bereavement support by a healthcare or other care professional (28), suggest that important opportunities for providing families with information about child grief and support services may be being missed (23).

### Strengths, weaknesses and implications for research

This free-text data collected from parent/caregiver participants in a large national study of people’s experiences of bereavement during the Covid-19 pandemic in the UK provides important insights into the difficulties, support needs and experiences of bereaved children and young people, from the perspective of their parents/caregivers. Limitations include the fact that we did not hear from children and young people directly, and that this data was collected from a single question which formed part of a broader questionnaire and study focused on adult experiences and needs. Lack of random sampling also means that the survey is not statistically representative of the whole bereaved population, and despite significant efforts and targeted recruitment, people from minority ethnic backgrounds and men are underrepresented in the data set overall and in responses to this question. Most bereavements were of grandparents, with small group sizes for other types of bereavements including those likely to have greater impact, such as death of a parent or sibling, hence needs for support may be greater than reported here. Future research is needed which focuses specifically on the experiences of children and young people, in particular from those population groups worst affected by Covid-19 in the UK, including Black and minority ethnic communities and those living in areas of high socio-economic deprivation. Research exploring school and specialist provider perspectives and experiences of supporting bereaved families during the pandemic, and in particular the use and acceptability of remote/on-line interventions, is also recommended.

### Conclusions and implications for policy and practice

These results demonstrate the added challenges faced by children and young people who experienced the death of a family member during the Covid-19 pandemic. These included loss-related issues caused by being unable to spend time with their relatives before they died as well as the disruption to usual coping mechanisms and resources caused by social isolation and school and university closures. Despite these difficulties, many parents felt that their children were resilient and coping well with their support, with the informal support of teachers, schools and wider family also valued. However, a significant minority described the additional, more specialist emotional or mental health support needed or received by their children. Barriers such as waiting lists and lack of appropriate support and information on how to get support prevented some families from accessing the support their children needed. Based on these study findings, we make three recommendations for improving the support available for bereaved children and young people:

1. Facilitating open communication and healthy grieving within families and among friends, via the promotion of age-appropriate, accessible self-help resources and materials made available online and via community organisations such as schools, libraries, GP practices and pharmacies. Provision of informal sessions aimed at improving community grief literacy with respect to children as well as adults, with specific consideration to the pandemic context in shaping grief experiences, is also recommended (9,12,33,34).
2. Training for school staff to have age-appropriate conversations with students around grief and bereavement, and to be able to identify when a child might need additional specialist support (which should also be available within the school setting). During periods of lockdown and school closure, regular check-ins with bereaved families are important. Specialist programmes should continue for existing students (remotely if needed), whilst also proactively identifying and engaging with newly bereaved families who may need support. Finding ways of facilitating peer support for older children may also help counter the effects of social isolation during lockdown periods.
3. Adequate resourcing of specialist childhood bereavement and mental health services to tackle gaps in some regions, reduce long waiting lists and enable longer term support, with children bereaved of a grandparent or other family member also considered eligible for intervention when needed. Information should be provided on available bereavement and mental health services by healthcare professionals following the death and made publicly available online, including NHS webpages, as well community settings such as libraries, schools and General Practice surgeries so that parents can more easily identify appropriate locally available support. Greater system-level investment and disaster-planning in child and adolescent mental health infrastructure is also needed to ensure the future resilience and adaptability of these urgent care services during pandemics.

## Data Availability

Full study data sets will be made available following study closure in February 2022. Data sharing requests will be considered prior to this and should be directed to Dr Emily Harrop

## Acknowledgements

Our thanks to everyone who completed the survey for sharing their experiences, and to all the individuals and organisations that helped disseminate the survey. We would also like to thank the project assistants, collaborators and study advisory group members.

## Author contributions

E.H. and L.E.S. designed the study, led the application for funding and are co-principal investigators; E.H. drafted the paper; M.L., A.B., D.F., A.N., A.T.B., A.P. are members of the research team or the study advisory group and contributed to the design of the study and survey. E.H. and S.G. conducted the thematic analysis of qualitative data. All authors contributed to drafting the paper and read and approved the final manuscript.

## Ethical approval

The study protocol and supporting documentation was approved by Cardiff University School of Medicine Research Ethics Committee (SMREC 20/59). The study was conducted in accordance with the Declaration of Helsinki and all respondents provided informed consent.

