## Supplementary file 1 for "Parental perspectives on the grief and support needs of children and young people bereaved during the Covid-19 pandemic: Qualitative findings from a national survey"

Thank you once again for taking part in our study and agreeing to be sent this second questionnaire. We really appreciate the time you have taken to help us with this study.

In this second questionnaire we are interested to find out more about your grief experiences and wellbeing at this point in time. We would also like to find out about any support that you have been using recently or feel like you may need and any difficulties you may be experiencing accessing support.

More detailed information about the study is included in the document *Information for Participants – Second Survey* that is enclosed in your survey letter. Please read this information to help you decide whether you would like to take part in this second survey and complete the consent section below if you would like to continue.

Information about bereavement support services and resources is also provided in the same document and at the end of this survey.

**Consent**

By participating in this survey, you agree that you have read and understood the information provided above and that you are aged 18 or over.

**I confirm that I have read and understood the information provided about the purpose of this study and how my data will be used. I agree to take part in the following survey knowing that all questions are optional and I can finish the survey at any point.**

Please tick the box to agree with the above statement

**Thank you for your help!**

**Before we start, could you please tell us whether you have experienced any other bereavements of close friends or family members since completing your last questionnaire two months ago?**

| Yes |
| --- |
| No |

**If you have been bereaved again, we are deeply sorry to hear this and appreciate that you may not feel like completing this survey right now. If you would prefer for us to contact you in six months instead, please let us know below and use the enclosed prepaid envelope to return your survey to us.**

| I would prefer to complete this survey another time. |
| --- |
| I would like to continue with the survey. |

**Comments:**

**If you would like to continue, please carry on with the survey.**

**Part A**

This first section contains a series of questions which will help us to understand how your grief is affecting you and how you are coping with and adjusting to your bereavement. Some of these questions you may remember from answering previously, whilst others are new to this questionnaire. For all questions please remember that there are no right or wrong answers.

**A1. To help us better understand how you have been adjusting to the loss of your [insert] and how things may have changed for you since your last questionnaire, please indicate your response to the following attitudes:**

|  | **Strongly agree** | **Agree** | **Neither agree nor disagree** | **Disagree** | **Strongly disagree** |
| --- | --- | --- | --- | --- | --- |
| 1. I feel able to face the pain which comes with loss. | 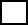 | 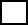 | 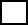 | 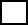 | 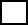 |
| 2. For me, it is difficult to switch off thoughts about the person I have lost. | 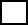 | 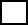 | 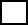 | 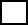 | 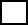 |
| 3. I feel very aware of my inner strength when faced with grief. | 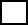 | 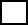 | 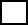 | 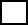 | 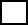 |
| 4. I believe that I must be brave in the face of loss. | 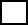 | 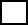 | 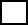 | 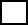 | 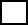 |
| 5. I feel that I will always carry the pain of grief with me. | 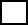 | 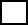 | 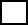 | 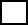 | 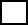 |
| 6. For me, it is important to keep my grief under control. | 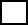 | 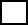 | 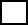 | 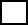 | 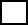 |
| 7. Life has less meaning for me after this loss. | 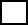 | 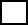 | 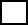 | 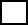 | 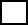 |
| 8. I think it’s best just to get on with life in spite of this loss. | 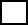 | 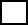 | 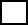 | 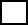 | 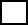 |
| 9. It may not always feel like it but I do believe that I will come through this experience of grief. | 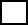 | 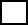 | 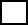 | 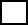 | 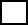 |

**Adult Attitude to Grief scale © Linda Machin 2000**

**A2. Several grief-reactions are listed below. Please indicate how often you have experienced each reaction in the past month, in response to the death of your [insert]:**

|  | **Never** | **Rarely** | **Sometimes** | **Frequently** | **Always** |
| --- | --- | --- | --- | --- | --- |
| 1. I had intrusive thoughts and images  related to the person who died. | 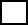 | 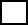 | 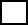 | 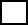 | 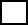 |
| 2. I experienced intense emotional pain,  sadness, or pangs of grief. | 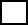 | 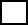 | 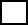 | 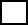 | 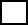 |
| 3. I found myself longing or yearning for the person who died. | 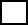 | 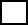 | 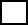 | 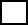 | 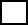 |
| 4. I experienced confusion about my role in life or a diminished sense of self. | 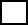 | 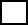 | 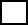 | 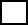 | 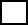 |
| 5. I had trouble accepting the loss. | 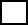 | 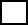 | 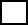 | 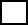 | 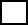 |
| 6. I avoided places, objects or thoughts  that reminded me that the person I lost has died. | 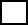 | 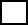 | 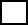 | 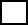 | 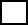 |
| 7. It was hard for me to trust others. | 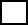 | 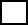 | 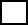 | 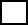 | 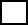 |
| 8. I felt bitterness or anger related to his/her death. | 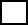 | 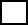 | 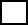 | 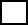 | 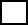 |
| 9. I felt that moving on (e.g. making new friends, pursuing new interests) was difficult for me. | 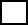 | 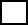 | 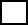 | 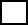 | 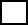 |
| 10. I felt emotionally numb. | 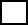 | 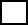 | 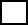 | 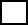 | 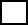 |
| 11. I felt that life is unfulfilling or meaningless without him/her. | 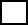 | 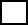 | 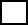 | 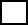 | 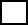 |
| 12. I felt stunned, shocked or dazed by his/her death. |  |  |  |  |  |
| 13. I noticed significant reduction in social, occupational, or other important areas of functioning (e.g., domestic responsibilities) as a result of his/her death. |  |  |  |  |  |
|  | **Never** | **Rarely** | **Sometimes** | **Frequently** | **Always** |
| 14. I had intrusive thoughts and images  associated with the circumstances of his/  her death. |  |  |  |  |  |
| 15. I experienced difficulty with positive reminiscing about the lost person. |  |  |  |  |  |
| 16. I had negative thoughts about myself in relation to the loss (e.g., thoughts about self-blame) |  |  |  |  |  |
| 17. I had a desire to die in order to be with the deceased. |  |  |  |  |  |
| 18. I felt alone or detached from other individuals. |  |  |  |  |  |

**Traumatic Grief Inventory**

**A3. We would also like to get a sense of how well supported you feel at the moment. Using the scale shown below please select the statement that best describes the way you have been feeling during the past two weeks, including today.**

|  | **Does not describe me at all** | **Does not quite describe me** | **Describes me fairly well** | **Describes me well** | **Describes me very well** |
| --- | --- | --- | --- | --- | --- |
| People take the time to listen to how I feel |  |  |  |  |  |
| I can express my feelings about my grief openly and honestly |  |  |  |  |  |
| It helps me to talk with someone who is non-judgemental about how I grieve |  |  |  |  |  |
| There is at least one person I can talk to about my grief. |  |  |  |  |  |
| I can get help for my grieving when I need it. |  |  |  |  |  |

**Inventory for Social Support (Hogan and Schmidt 2002)**

**A4. Next we would like to ask you four questions about your feelings on aspects of your life. For each of these questions we would like you to give an answer on a scale of 0 to 10, where 0 is “not at all” and 10 is “completely”.**

|  | **Please rate each question on a scale from 0 (‘not at all’) to 10 (‘completely’)** | | | | | | | | | | |
| --- | --- | --- | --- | --- | --- | --- | --- | --- | --- | --- | --- |
|  | **0** | **1** | **2** | **3** | **4** | **5** | **6** | **7** | **8** | **9** | **10** |
| Overall, how satisfied are you with your life nowadays? |  |  |  |  |  |  |  |  |  |  |  |
| Overall, to what extent do you feel that the things you do in your life are worthwhile? |  |  |  |  |  |  |  |  |  |  |  |
| Overall, how happy did you feel yesterday? |  |  |  |  |  |  |  |  |  |  |  |
| On a scale where 0 is “not at all anxious” and 10 is “completely anxious”, overall, how anxious did you feel yesterday? |  |  |  |  |  |  |  |  |  |  |  |

**Source: Office National Statistics wellbeing measure**

**Part B. Bereavement support**

**This section includes questions on your experiences of accessing support and help with your bereavement over the last two months.**

**B1. Over the last two months have you experienced any difficulties getting support for your grief and bereavement?**

|  | Yes | Somewhat | No | I’ve not tried to get their support |
| --- | --- | --- | --- | --- |
| From friends and family |  |  |  |  |
| From GP surgery |  |  |  |  |
| From bereavement services |  |  |  |  |

**B2. Do any of the responses below describe your experiences over the last two months? (Please select all that apply):**

| I have not wanted any support from bereavement services because my family and friends provide me with enough support |
| --- |
| I have not wanted any support from bereavement services because I am coping ok without this type of support |
| I have not wanted any support from bereavement services because I do not think it would help me |
| I do not know how to get support from bereavement services |
| I have felt uncomfortable asking for support from bereavement services |
| I have felt uncomfortable asking for help or support from friends or family |
| The support I wanted from bereavement services was not available to me |
| Friends or family have not been able to support me in the way I that wanted |

**B3. Please describe any difficulties that you have faced getting support from either friends/family or bereavement services.**

**Section C. Types of support used**

**This section includes questions on the types of support you have been using since becoming bereaved. It includes more detailed questions about support that you may previously have told us about and asks about any new support that you may have been receiving since your last questionnaire.**

**C1. In your previous questionnaire you told us that you [personalise]. We would like to know a bit more about this support. Firstly, are you still using this support?**

| **Yes**  *If yes, please answer questions a-g below.* |
| --- |
| **No**  *If no, please tell us when you stopped using this support and then answer questions a-g below.*  Support no longer used from: …………………………………………… |

**Can you please also tell us:**

| a) How is/was the support provided? (e.g. in person, over the phone, via skype/zoom, web-chat etc) |
| --- |
| b) How effective do (or did) you find this method of communication? |
| c) Roughly when did you first start using this support? |
| d) How often do/did you use this support? If relevant please state the number of sessions received. |
| e) Do you feel that you started using this support at around the right time for you? |
| f) Do you feel happy with the amount of support you have received from this support provider? |
| g) How did you first find out about this support? |

**You also told us you had been getting support [personalise]. Are you still receiving this support?**

| **Yes**  *If yes, please answer questions a-g below.* |
| --- |
| **No**  *If no, please tell us when it finished and then answer questions a-g below.*  Support no longer received/used from: …………………………………………… |

**Please can you also tell us:**

| a) How is/was the support provided? (e.g. in person, over the phone, via skype/zoom, web-chat etc) |
| --- |
| b) How effective do (or did) you find this method of communication? |
| c) Roughly when did you first start using this support? |
| d) How often do/did you use this support? If relevant please state the number of sessions received. |
| e) Do you feel that you started using this support at around the right time for you? |
| f) Do you feel happy with the amount of support you have received from this support provider? |
| g) How did you first find out about this support? |

**Have you used any other bereavement support services or groups since completing your last survey two months ago?**

| **Yes**  *If yes, please continue to the* ***next question (C2)*** *below.* |
| --- |
| **No**  *If no, please continue to* ***Section D on page 18.*** |

**C2. If you indicated above that you have also used some other support groups, services or resources, please tell us what type of support this is:**

| Telephone helpline support (e.g. bereavement helpline) |
| --- |
| Online community support via written comments (e.g. Facebook group, online chat forum) |
| Informal support/talking group (e.g. social group for bereaved people) |
| Formal bereavement support group (e.g. group discussions about bereavement guided by a facilitator; or group counselling) |
| One-to-one support (e.g. individual counselling) |
| Specialist mental health support |
| Other |

**If you selected Other, please specify:**

________________________________________________________________________

**Are you still receiving this support?**

| **Yes**  *If yes, please answer questions a-h below.* |
| --- |
| **No**  *If no, please tell us when it finished and then answer questions a-h below.*  Support no longer received/used from: …………………………………………… |

**Please can you also tell us the following details:**

| a) Who provides/provided this support? (e.g. name of organisation or group) |
| --- |
| b) How is/was the support provided? (e.g. in person, over the phone, via skype/zoom, web-chat etc) |
| c) How effective do (or did) you find this method of communication? |
| d) Roughly when did you first start using this support? |
| e) How often do/did you use this support? If relevant, please state the number of sessions received. |
| f) Do you feel that you started using this support at around the right time for you? |
| g) Do you feel happy with the amount of support you have received from this support provider? |
| h) How did you first find out about this support? |

**Have you used any other additional bereavement support services, groups or resources since completing your last survey two months ago?**

| **Yes**  *If yes, please continue to the* ***next question (C3)*** *below.* |
| --- |
| **No**  *If no, please* ***continue to Section D on page 18****.* |

**C3. If you indicated above that you have also used some other additional support groups, services or resources, please tell us what type of support this is:**

| Telephone helpline support (e.g. bereavement helpline) |
| --- |
| Online community support via written comments (e.g. Facebook group, online chat forum) |
| Informal support/talking group (e.g. social group for bereaved people) |
| Formal bereavement support group (e.g. group discussions about bereavement guided by a facilitator; or group counselling) |
| One-to-one support (e.g. individual counselling) |
| Specialist mental health support |
| Other |

**If you selected Other, please specify:**

________________________________________________________________________

**Are you still receiving/using this support?**

| **Yes**  *If yes, please answer questions a-h below.* |
| --- |
| **No**  *If no, please tell us when it finished and then answer questions a-h below.*  Support no longer received/used from: …………………………………………… |

**Please can you also tell us the following details:**

| a) Who provides/provided this support? (e.g. name of organisation or group) |
| --- |
| b) How is/was the support provided? (e.g. in person, over the phone, via skype/zoom, web-chat etc) |
| c) How effective do (or did) you find this method of communication? |
| d) Roughly when did you first start using this support? |
| e) How often do/did you use this support? If relevant, please state the number of sessions received. |
| f) Do you feel that you started using this support at around the right time for you? |
| g) Do you feel happy with the amount of support you have received from this support provider? |
| h) How did you first find out about this support? |

**In addition to the two further sources of support you have just described, are there any other bereavement support services, groups or resources that you have used in the two months since completing your last survey?**

| **Yes**  *If yes, please continue to the* ***next question (C4)*** *below.* |
| --- |
| **No**  *If no, please* ***continue to Section D on page 18****.* |

**C4. If you indicated above that you have also used a third form of additional support, please tell us what type of support this is:**

| Telephone helpline support (e.g. bereavement helpline) |
| --- |
| Online community support via written comments (e.g. Facebook group, online chat forum) |
| Informal support/talking group (e.g. social group for bereaved people) |
| Formal bereavement support group (e.g. group discussions about bereavement guided by a facilitator; or group counselling) |
| One-to-one support (e.g. individual counselling) |
| Specialist mental health support |
| Other |

**If you selected Other, please specify:**

________________________________________________________________________

**Are you still receiving/using this support?**

| **Yes**  *If yes, please answer questions a-h below.* |
| --- |
| **No**  *If no, please tell us when it finished and then answer questions a-h below.*  Support no longer received/used from: …………………………………………… |

**Please can you also tell us the following details:**

| a) Who provides/provided this support? (e.g. name of organisation or group) |
| --- |
| b) How is/was the support provided? (e.g. in person, over the phone, via skype/zoom, web-chat etc) |
| c) How effective do (or did) you find this method of communication? |
| d) Roughly when did you first start using this support? |
| e) How often do/did you use this support? If relevant, please state the number of sessions received. |
| f) Do you feel that you started using this support at around the right time for you? |
| g) Do you feel happy with the amount of support you have received from this support provider? |
| h) How did you first find out about this support? |

**Section D. Ongoing Support Needs**

**This section includes questions on the kinds of support or help that you may have needed over the last two months. It also asks how well you feel these needs have been met by friends/family and any other sources of support you may have been using.**

**D1a. Over the last two months, please tell us how much support or help you have needed with the following:**

| **Practical tasks** | **High level of support needed** | **Fairly high level of support needed** | **Moderate level of support needed** | **Little support needed** | **No support needed** |
| --- | --- | --- | --- | --- | --- |
| Practical and administrative tasks e.g. finances, other paperwork etc. |  |  |  |  |  |
| Getting relevant information and advice e.g. legal, financial, available support |  |  |  |  |  |
| Looking after myself/family e.g. getting food, medication, childcare |  |  |  |  |  |

If you have needed support with the above (*practical tasks*), please tell us how helpful you have found the support of other people, groups or resources in meeting these needs:

| **Sources of support** | **Very helpful** | **Slightly helpful** | **Neither helpful nor unhelpful** | **Slightly unhelpful** | **Very unhelpful** |
| --- | --- | --- | --- | --- | --- |
| Friends and family |  |  |  |  |  |
| **[personalise]** |  |  |  |  |  |
| **[personalise]** |  |  |  |  |  |
| **Sources of support** | **Very helpful** | **Slightly helpful** | **Neither helpful nor unhelpful** | **Slightly unhelpful** | **Very unhelpful** |
| Other type of support 1  *if relevant, please state here:* |  |  |  |  |  |
| Other type of support 2 *if relevant, please state here:* |  |  |  |  |  |

Please explain some of the ways you have been helped with your practical support needs. Alternatively, you might like to tell us about any further help that you need with these.

**D2a. Over the last two months, please tell us how much support or help you have needed with the following:**

| **Managing my grief** | **High level of support needed** | **Fairly high level of support needed** | **Moderate level of support needed** | **Little support needed** | **No support needed** |
| --- | --- | --- | --- | --- | --- |
| Dealing with my feelings about being without my loved one |  |  |  |  |  |
| Dealing with my feelings about the way my loved one died |  |  |  |  |  |
| Expressing my feelings and feeling understood by others |  |  |  |  |  |
| Feeling comforted and reassured |  |  |  |  |  |

If you have needed support with the above (managing your grief), please tell us how helpful you have found the support of other people, groups or resources in meeting these needs:

| **Sources of support** | **Very helpful** | **Slightly helpful** | **Neither helpful nor unhelpful** | **Slightly unhelpful** | **Very unhelpful** |
| --- | --- | --- | --- | --- | --- |
| Friends and family |  |  |  |  |  |
| **[personalise]** |  |  |  |  |  |
| **[personalise]** |  |  |  |  |  |
| Other type of support 1  *if relevant, please state here:* |  |  |  |  |  |
| Other type of support 2 *if relevant, please state here:* |  |  |  |  |  |

Please explain some of the ways in which you have been helped with *managing your grief*. Alternatively, you might like to tell us about any further help that you need with this.

**D3a. Over the last two months, please tell us how much support or help you have needed with the following:**

| **Quality of life and mental wellbeing** | **High level of support needed** | **Fairly high level of support needed** | **Moderate level of support needed** | **Little support needed** | **No support needed** |
| --- | --- | --- | --- | --- | --- |
| Finding balance between grieving and other areas of life |  |  |  |  |  |
| Participating in work, leisure or other regular activities (e.g. shopping, housework) |  |  |  |  |  |
| Managing and maintaining my relationships with friends and family |  |  |  |  |  |
| Loneliness and social isolation |  |  |  |  |  |
| Feelings of anxiety and depression |  |  |  |  |  |
| Regaining sense of purpose and meaning in life |  |  |  |  |  |
| Feeling optimistic and hopeful for the future |  |  |  |  |  |

If you have needed support with the above (quality of life and mental wellbeing), please tell us how helpful you have found the support of other people, groups or resources in meeting these needs:

| **Sources of support** | **Very helpful** | **Slightly helpful** | **Neither helpful nor unhelpful** | **Slightly unhelpful** | **Very unhelpful** |
| --- | --- | --- | --- | --- | --- |
| Friends and family |  |  |  |  |  |
| **[personalise]** |  |  |  |  |  |
| **[personalise]** |  |  |  |  |  |
| Other type of support 1  *if relevant, please state here:* |  |  |  |  |  |
| Other type of support 2 *if relevant, please state here:* |  |  |  |  |  |

Please explain some of the ways in which you have been helped with your *quality of life and mental wellbeing.* Alternatively, you might like to tell us about any further help that you need with this.

**D4. Do you have any other needs for help or support that have not already been described?**

| Yes |
| --- |
| No |

If yes, please describe:

**D5. Are there any children or young people living with you who have also been affected by this bereavement?**

| Yes |
| --- |
| No |

If yes, how old are they?

……………………………………………………………………………………………………………………………………………….

**Please tell us about any support that you feel they need and/or any support they have been receiving:**

**D6. Do you feel that there are any changes to government policy or support services that could be made to help you at this time (and which you have not already described)?**

**D7. If you would like to write anything further about your experiences of grieving over the last few months and/or how you feel you have been coping please use the box below:**

**Section E.**

**In this section we would like to ask you a few additional questions about other aspects of your life that may affect or have been affected by your bereavement.**

**E1. Have you been diagnosed with any illness or medical condition since you completed the previous survey two months ago?**

| Yes |
| --- |
| No |

If yes, please provide details:

……………………………………………………………………………………………………………………………………………….

**E2. Roughly how many appointments with your GP have you had over the last two months?**

……………………………………………………………………………………………………………………………………………….

**E3. Roughly how many times have you bought over-the-counter medicine (e.g. paracetamol, sleep aids or stress relief remedies) over the last two months?**

| Never |
| --- |
| One to three times |
| 4 times or more |

**E4. Have you experienced any difficulties with sleeping over the last two months?**

| Yes |
| --- |
| No  *If no, please continue to question E5.* |

**If yes, how often do you have difficulties with sleeping (e.g. trouble falling asleep, waking up early or in the middle of the night, bad dreams, poor overall sleep quality)?**

| Less than once a week |
| --- |
| Once or twice a week |
| Three or more times a week |

**To enable us to better understand how bereavement impacts upon the employment and** **working life of bereaved people, please answer the following questions.**

**E5. Has your employment status changed since your last survey two months ago?**

| **Yes** |
| --- |
| **No** |
| **Not relevant for me**  *(e.g. retired, full-time student, permanently sick/disabled, long-term unemployed, looking after the home, caring for a loved one)*  **If not relevant for you, please continue to the end of the survey on page 28.** |

If yes, please provide details (including if you have been furloughed):

……………………………………………………………………………………………………………………………………………….

**E6. Approximately how much time have you had off work since becoming bereaved (in days/weeks or months)?**

| Amount of time off work due to bereavement, illness or stress: |
| --- |
| Amount of time off work due to furlough or unemployment: |
| Amount of time off work due to other extended periods of leave (please specify below): |

If you indicated above that you had time off work for other reasons (e.g. school holidays/closures, caring responsibilities, maternity leave), please specify here:

……………………………………………………………………………………………………………………………………………….

**E7. If you would like to share further information about your time off work since your bereavement, please use the box below:**

**Many thanks for completing this second survey and for all of the time that you have given to this research so far.** Your responses will help to enable others who are bereaved to access the support they need. We appreciate that this may have been difficult and painful for you and we are very grateful for your contribution.

We would like to send you one final questionnaire six months from now, which will enable us to consider the longer term impacts of bereavement during the Covid-19 pandemic. Please use the box below to tell us if you are happy for us to send you one final questionnaire.

| Yes |
| --- |
| No |

Thank you again for your help. We are extremely grateful for your contribution.

If you would like to talk to somebody about your bereavement, you can access support from these services:

- Marie Curie Bereavement Support: 0800 090 2309 <https://www.mariecurie.org.uk/help/support/bereaved-family-friends/dealing-grief/bereavement-or-grief-counselling>
- Cruse Bereavement Care: 0808 808 1677

<https://www.cruse.org.uk/>

- NHS Bereavement Helpline: 0800 2600 400 <https://www.nhs.uk/conditions/stress-anxiety-depression/coping-with-bereavement/>
- The Good Grief Trust: <https://www.thegoodgrieftrust.org/>
- At a Loss: [www.ataloss.org](http://www.ataloss.org)

***Thank you again for your help!***
